# Individual effect of diet on postprandial glycemic response and its relationship with gut microbiome profile in healthy subjects: protocol for a series of randomized N-of-1 trials

**DOI:** 10.1101/2023.12.22.23300429

**Authors:** C.P. Zamparette, B.L. Teixeira, G.N.F. Cruz, V. B. Filho, L.F.V. De Oliveira

## Abstract

**Background:** Type 2 diabetes causes over a million deaths annually, ranking in the top ten causes of death worldwide. Glycemic control through dietary adequacy is essential for treatment success and disease prevention. Recent evidence indicates that the glycemic response to various foods varies from individual to individual. The intestinal microbiome is seen as a potential key player, mediating the effect of foods on glycemic response. By design, however, most published studies cannot separate variation in the individual treatment effects (ITE) of different diets from within-individual variability of glycemic responses. In this context, the present study aims to assess the heterogeneity in the ITE of diet on glycemic response and investigate the relevance of the intestinal microbiome profile as a predictor of this heterogeneity.

**Methods:** This study is a series of N-of-1 randomized clinical trials. Each participant will undergo five treatment cycles of two prescribed diets (low-carb versus vegan) in one of two randomly chosen treatment sequences (ABBABAABBA or BAABABBAAB). The primary outcome is the positive incremental area-under-the-curve (iAUC) of the postprandial interstitial fluid glucose measured within 2 hours of meal consumption. The trial plans to recruit 80 healthy volunteers with ages between 18 and 60. Fecal samples will be collected at baseline for microbiome analysis by metagenomics shotgun technique. Random effects linear models will be used for the primary analysis.

**Discussion:** While significant variation of individual effects warrants personalized interventions, it is well-known that glycemic responses to the same food, in the same individual, vary from occasion to occasion. Yet, most clinical studies are based on designs that are incapable of separating ITE variation from within-individual variability. This is a major limitation since the personalization of dietary interventions is only justified by clinically relevant heterogeneity of individual-level effects. In this study, if significant ITE variation is indeed observed, then we will also be able to estimate the relationship between the intestinal microbiome and the expected diet effects. This is essential to identify predictive biomarkers, which can identify those who intrinsically benefit the most from which diet, going beyond pure associations with glycemic response. Conversely, observing negligible ITE variation in a large series of N-of-1 trials would cast major doubts on the relevance of personalizing dietary interventions for glycemic control. Therefore, the present study represents a major step toward understanding the clinical value of microbiome-driven precision nutrition.

## Introduction

Non-communicable diseases (NCDs) are globally widespread, with cardiovascular diseases, cancer, diabetes, and chronic respiratory diseases being major contributors. These NCDs account for approximately three-quarters of annual global deaths ^1^, equating to a person dying every 2 seconds due to these diseases. NCDs are a critical issue affecting all countries and regions, particularly developing and underdeveloped nations, where about 86% of these deaths occur in individuals under 70 years old ^2^. Most of the deaths and health problems caused by NCDs could be prevented or delayed by addressing behavioral risk factors such as smoking, unhealthy diet, harmful alcohol use, sedentary lifestyle, and cardiometabolic risk factors like high blood pressure, overweight, obesity, hypercholesterolemia, and hyperglycemia ^2^.

Diabetes mellitus, a severe chronic metabolic disorder characterized by high blood glucose levels ^3^, is a growing global health emergency. In 2021, approximately half a billion people worldwide had diabetes, projected to rise to around 643 million by 2030 and 783 million by 2045 (IDF DIABETES ATLAS, 2022). Type 2 diabetes (DM2) is the prevalent form, comprising over 95% of all diabetes cases ^4^. Long-term insulin deficiency or insensitivity can lead to serious complications, including cardiovascular diseases, neuropathy, nephropathy, lower limb amputation, and retinopathy ^5^.

Diet and nutrition are crucial for managing diabetes, particularly postprandial glycemic control ^6^. Glycemic response to food intake depends on carbohydrates, starch nature, fat and protein content, fiber, particle size, food processing, and ingestion method ^7,8^. Postprandial glycemia is associated with type 2 diabetes development, cardiovascular diseases, and mortality ^9,10^.

In recent research, the intestinal microbiome has gained attention as a potential key player in nutritional contexts and glycemic control ^11^. With over a thousand species, the gut microbiome influences homeostasis, circadian rhythms, nutritional and metabolic responses, pathogen protection, and immunomodulation ^12,13^. Dysbiosis, an imbalance in the gut microbiome, is linked to various cardiovascular, neurological, immune, and metabolic pathologies. These disruptions impact liver-bile metabolism, and promote inflammation, insulin resistance, and incretin secretion, potentially leading to obesity, metabolic syndrome, and type 2 diabetes ^14–16^.

Additionally, dysbiosis is believed to contribute to reduced beneficial microorganism abundance and signaling, facilitating colonization by harmful microbes ^17^. Dysbiosis could link chronic inflammation and gastrointestinal diseases through various molecular pathways, including TGF-β, NFκB, TNF-α, and ROS. Uncontrolled chronic inflammation increases the risk of type 2 diabetes, irritable bowel syndrome, and colorectal cancer ^18^. However, the gut microbiome’s diverse composition and functions vary even among healthy individuals due to genetics, epigenetics, and lifestyle factors such as diet ^19,20^. The gut microbiome produces numerous metabolites capable of systemic metabolic and immunological pathway modulation. Short-chain fatty acids (SCFAs) impact glucose homeostasis and adipose tissue inflammation ^21,22^.

Considering the potential link between the microbiome and glycemic response, a series of N-of-1 trials is proposed to evaluate individual diet effects on postprandial glycemic response and its correlation with the intestinal microbiome profile. The study aims to determine the presence of individual diet effect heterogeneity and its correlation with microbiome characteristics, contributing to personalized nutritional interventions for glycemic control based on the microbiome.

### Objectives

The specific objectives were categorized as primary and secondary. The primary objectives follow pre-specified analyses in the protocol, while the secondary objectives involve exploratory analyses for hypothesis generation.

1. Primary Objectives

A. Determining the presence of heterogeneity in the individual treatment effects of diet on postprandial glycemic response.
B. Determine the influence of the intestinal microbiome profile on the variability of individual treatment effects of diet on postprandial glycemic response.
2. Secondary Objectives

A. Explore associations between individual treatment effects of diet and intestinal microbiome markers.
B. Explore associations between postprandial glycemic response itself and intestinal microbiome markers.

### Trial design

The study is a series of N-of-1 trials randomized into two treatment sequences in a 1:1 allocation ratio. The major objective of the study is to assess the heterogeneity of the individual diet effects on postprandial glycemic response and its relationship with gut microbiome profile. To achieve this, participants will be provided with two breakfast meals, one with a low carbohydrate content and the other vegan, which they will consume for ten consecutive days, alternating between the meals according to the treatment sequence they are assigned to.

During the intervention days, participants will use a continuous intradermal glucose monitoring device and measure fasting glucose levels and postprandial glucose every 30 minutes until two hours have elapsed after consuming breakfast. Throughout this period, participants should not eat any other type of food (other than water) nor should they perform any physical activity. Additionally, to assess dietary consumption beyond breakfast, participants will be instructed to record the foods they consume throughout the day.

Before dietary intervention starts, participants will be required to provide a stool sample for two purposes: one for gut microbiome analysis and the other for the isolation and characterization of fecal bacteria.

## Methods: Participants, interventions, and outcomes

### Study setting

The recruitment of 80 participants, as well as the analysis of samples, will be carried out at BiomeHub, located in Florianopolis, Santa Catarina, Brazil.

### Eligibility criteria

Eighty healthy volunteers will be recruited to assess postprandial glycemia in response to a standardized meal.

- Inclusion Criteria

○ Healthy men or women aged 18 to 60.
○ BMI > 18.5 and < 30.
○ Willing to use an intradermal continuous glucose monitoring sensor during the 10-day study.
○ Own a mobile phone with NFC technology.
○ Willing to provide a fecal swab sample and a stool sample.
○ Understand, agree, and sign the Informed Consent Form (ICF) approved by the Ethics Committee (CEP).
- Exclusion Criteria

○ Pregnant or lactating women.
○ Diagnosis of any gastrointestinal disorder or disease (Irritable Bowel Syndrome, Ulcerative Colitis, Crohn’s Disease).
○ Intolerance or allergy to any diet ingredient.
○ Autoimmune disorder (Lupus, Type 1 Diabetes, Celiac Disease) or infectious disease.
○ Diabetes diagnosis.
○ Cancer diagnosis, acute myocardial infarction, or stroke in the last 6 months.
○ Use of hypoglycemic medication.
○ Use of proton pump inhibitors, immunosuppressants, or antimicrobials in the last 3 months.
○ Use of laxative medications in the last 30 days.
○ Underwent invasive procedures or surgery in the last 6 months.
○ Admission to ICU in the last 2 years.
○ Participation in any experimental study or ingestion of any experimental drug within twelve months before the start of this study, by RDC 251/97.
○ Inability to read and understand the Informed Consent Form.

### Who will take informed consent?

Potential study participants will be screened by a research team member, which will involve a brief interview with the participant to assess eligibility for inclusion. Trained researchers will elucidate the primary study objectives, the criteria for inclusion and exclusion, as well as the interventions to be carried out. All study-related information will be provided and remain accessible to participants for consultation. Volunteers can address their inquiries during the interview with the researchers or subsequently via email. The participant consent document and personal survey will be collected during the interview or remotely through Google Forms. Participants will receive a copy of the signed consent form. The right of a participant to refuse participation without giving reasons will be respected.

### Additional consent provisions for collection and use of participant data and biological specimens

Additional consent provisions are not anticipated. All participants’ information will remain confidential and will solely be utilized for this research. Study findings will be published without any details that could disclose the volunteer’s identity. The biological specimens will be exclusively employed for this research and will be discarded after all necessary tests have been conducted.

## Interventions

### Explanation for the choice of comparators

The dietary interventions comprise two breakfast meals, based on diets typically recognized as healthy and used in Diabetes Mellitus control, which consist of a low-carbohydrate diet and a vegan diet ^23^. Both meals will adhere to dietary patterns featuring foods commonly consumed in the Brazilian diet, as outlined in the Brazilian Dietary Guidelines for the Population (BRASIL, 2014). They will be composed of a low-carbohydrate cake containing animal-derived ingredients and a cake with recommended carbohydrate quantities containing plant-based ingredients, including fruit in its composition. One of the meals will have a glycemic control characteristic, with a low carbohydrate content, below 45% of the carbohydrates recommended by the Acceptable Macronutrient Distribution Ranges (AMDR), in contrast, the other meal will consist entirely of plant-based foods in the recommended macronutrient proportions (Padovani et al., 2006). They will be similar in calories and standardized at 25% of daily calories, equivalent to the caloric distribution of breakfast, calculated by gender using the Protocol’s formulas - Harris & Benedict for individuals aged 15 to 74 ^24^. Each meal will consist of a cake/muffin, with approximately 100g and around 450 Kcal. The low-carbohydrate cake will contain banana, egg, almond flour, cocoa powder, stevia sweetener, and baking powder. The vegan cake will contain banana, wheat flour, cocoa powder, sugar, shredded coconut, canola oil, soy milk, and baking powder. Additionally, participants can choose to have unsweetened black coffee or water, which are beverages that won’t influence glycemic response.

### Intervention description

Each participant will undergo five cycles of two standardized meals to be administered during breakfast on two consecutive days. The meals will be prepared by an accredited nutritionist. The order of meals for each participant will follow one of two doubly counterbalanced sequences determined by 1:1 randomization (i.e., half of the participants ABBABAABBA, and the other half BAABABBAAB). Since each cycle consists of two breakfasts, each participant will be subjected to 10 days of intervention.

### Criteria for discontinuing or modifying allocated interventions

Participants can withdraw their consent to participate in the study at any time, without the obligation to justify their decision and without further consequences. The research team may recommend the withdrawal of participants from the intervention if it is deemed to be in the best interest of the participant. Participants who withdraw from the study will not be replaced.

### Strategies to improve adherence to interventions

Personally, during the dispensing of collection kits, dietary interventions, and glucose measurement devices, a conversation will take place emphasizing the importance of following study guidelines and adhering to the daily diet accurately. Instructions will be provided on when and under what conditions the foods should be consumed, how to use and care for the glucose measurement device, how to record foods consumed during the study, and how to collect fecal samples. The need for and the importance of contacting the researchers in case of any issues related to the study or missed food consumption will also be emphasized. To promote adherence, participants will receive protocol reminders via text messages, and they will be encouraged to reach out to the study staff with any concerns through phone or email. To assess adherence, electronic data collected from glucose monitoring will be reviewed, along with the completion of the daily food consumption record.

### Relevant concomitant care permitted or prohibited during the trial

Before breakfast, participants will measure their fasting interstitial fluid glucose, and consuming this meal, they will measure their postprandial interstitial fluid glucose every 30 minutes until the two-hour period, so, there will be 4 glucose measurements after the meal. During this two-hour interval, participants should abstain from consuming any other food or drink, except for water. Furthermore, engaging in physical exercise before the fasting interstitial fluid glucose measurement or after breakfast but before the postprandial interstitial fluid glucose measurement (until 2 hours after food ingestion) is not recommended.

### Provisions for post-trial care

Participants will receive compensation and full, immediate, and free assistance for as long as necessary in the event of damages or injuries resulting from participation in this study.

### Outcomes

The primary outcome is the positive incremental area-under-the-curve (iAUC) of the postprandial interstitial fluid glucose measured within 2 hours of meal consumption.

## Participant timeline

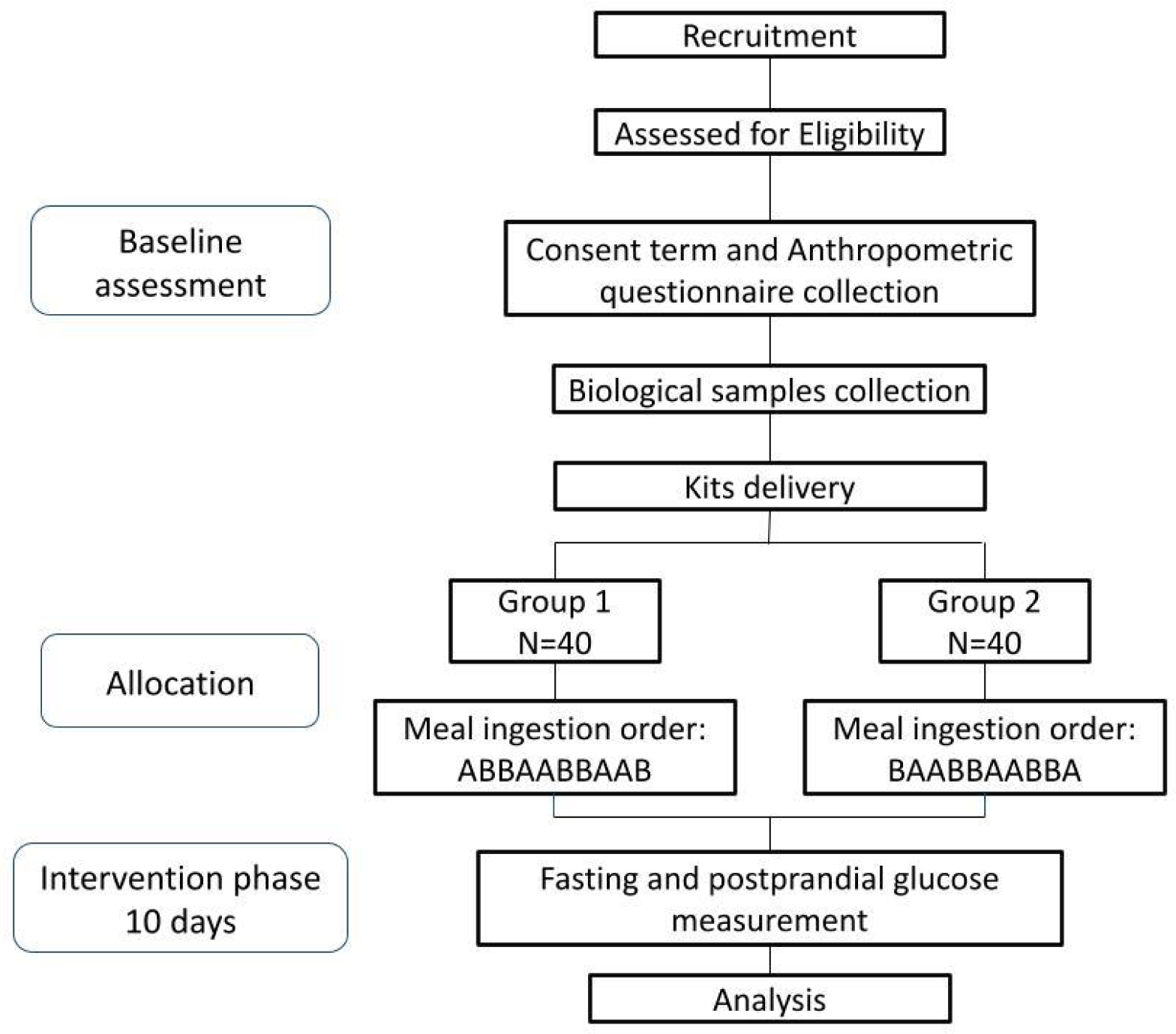

### Sample size

The sample size was calculated using simulation-based power calculation, assuming a Gaussian random effects model comparing two diets, with random intercepts and slopes varying per participant. A minimum clinically significant difference (MCID) of 40 mmol min L^-1^ was determined. The MCID value was used to define the narrowest range of individual treatment effects with clinical importance, which then informed simulation parameters. Using likelihood ratio tests at a 5% significance level and assuming a dropout rate of 20%, the present study required the recruitment of 80 participants followed through five treatment cycles. The number of participants and treatment cycles were selected to provide a statistical power of at least 90% for the detection of patient-by-treatment interaction (primary objective A) and of at least 85% for the detection of microbiome-by-treatment interaction (primary objective B). We adjusted p-values for multiple comparisons using Holm’s method. All details and simulation assumptions are available in the Statistical Analysis Plan (Supplementary data). The simulation code is fully reproducible and available at https://github.com/biomehub/bhub-n-of-1-sap.

### Recruitment

Study information will be disseminated through videos, images, and text posting on social media platforms like Facebook, Instagram, and LinkedIn, where participants will also be recruited. Interested individuals will be forwarded through a link that will lead to a document with several screening questions about the inclusion and exclusion criteria of the study. This information will be reviewed by responsible researchers who will select the participants. Eligible participants will be notified via email and will receive an electronic form of consent, an anthropometric questionnaire, study protocol details, kit collection, and biological sample delivery sites. Volunteers can address any questions or concerns by sending an email, WhatsApp message, or scheduling a call with the researchers. Once the consent term and the questionnaire are assigned, participants will receive the sample collection kit at their provided address. After biological sample collection, participants must deliver them to delivery locations where they will receive the intervention kits containing the standardized foods and a glucose measuring device. Ineligible volunteers or those placed on a waitlist will receive an email notification within 5 business days. Individuals who agree to participate in the study will receive a copy of the consent form via email along with comprehensive information about the study interventions. Furthermore, volunteers can choose a start date for the study from the available options. After selecting a date, participants must visit one of the kit delivery locations.

### Assignment of interventions: allocation

Each participant will be submitted to ten days of intervention, divided into five cycles of two standardized breakfasts on two consecutive days. The low-carb muffin will be designated as “A”, and the vegan muffin will be designated as “B”. Participants will be allocated into one of two doubly counterbalanced sequences, ABBABAABBA or BAABABBAAB, in a 1:1 allocation ratio using block randomization.

### Sequence generation

Upon enrollment, participants will be randomized into one of two doubly counterbalanced treatment sequences (ABBABAABBA or BAABABBAAB). The randomization schedule will be generated by the trial statistician (GNFC) who is not involved in the operational aspects of the study.

### Concealment mechanism

Randomization schedule will be prepared by the trial statistician (GNFC) who is not involved in the operational aspects of the study. The researchers will only become aware of each participant’s assigned sequence at the time of kit delivery for the intervention. This approach helps ensure that the randomization process remains unbiased and that researchers cannot influence sequence assignments.

### Implementation

The statistician responsible for the statistical data analysis will be in charge of generating the allocation sequence of the participants. Since the intervention consists of a standardized meal, and the groups only differ in the order in which these meals will be consumed, the researchers in charge of recruiting and monitoring the participants will handle the allocation of them.

### Assignment of interventions: Blinding Who will be blinded

This study will be conducted in an open-label manner, where the participants, investigators, site study teams, and project management teams will have knowledge of the allocation of interventions. Reasons supporting the choice of an open-label design included the study’s primary outcome (glycemic response), which is regarded as an objective and non-subjective endpoint.

### Procedure for unblinding if needed

Because this is an open-label study, there will be no need for an unblinding procedure.

## Data collection and management

### Plans for assessment and collection of outcomes

All study information will be provided to participants online. To assess volunteer eligibility, the candidates will be referred to an online questionnaire, via Google Forms, that covers the inclusion and exclusion criteria. Eligible participants will sign the online consent term and an anthropometric questionnaire, also via Google Forms, in which most of the questions will be multiple-choice to facilitate participant engagement and, subsequently, the analysis of results. In addition, participants must fill out a diet diary documenting their food intake throughout the intervention. Furthermore, participants will use the FreeStyle Libre app to record subcutaneous interstitial fluid glucose measurements. This app was selected because it is part of the subcutaneous interstitial fluid glucose measurement system included in the study package and, in addition, the researchers can monitor the measurements once the participants share the data with them.

### Plans to promote participant retention and complete follow-up

After participants have submitted their biological samples and received the study kits, during the 10-day intervention period, they will receive WhatsApp messages reminding them to measure fasting and post-prandial interstitial fluid glucose levels and to follow the specified meal order. Furthermore, every two days, participants will receive a message inquiring about any questions they may have, whether they are finding it easy to adhere to the interventions, if they are encountering any difficulties, and to remind them to complete the dietary diary. This will help us gain a better understanding of their daily routines. Nevertheless, rates of withdrawal and premature intervention discontinuation will be monitored. In the event that these rates exceed our expectations, a strategy may be required to ensure study power is maintained. If applicable, any changes to the protocol will be submitted for regulatory approval.

### Data management

All participant data will be entered electronically, a task to be performed by investigators. Original study forms will be entered and retained in a designated folder for each participant. Physical forms will be scanned and stored in the same folder. Participant files will be organized in numerical order and securely kept in an accessible location. These participant files will be maintained in storage for a period of 5 years following the study’s completion, in compliance with prevailing laws and regulations. Access to these documents will be restricted to the principal investigators and the responsible data analysis.

All physical consent forms and any other documents related to study data will be securely stored in locked cabinets.

### Confidentiality

Any participant data held by the company is stored with strict security standards, including protection against unauthorized access to their systems, restricted access by specific individuals to the location where personal information is stored, database encryption at rest, and authentication of access attempts.

### Plans for collection, laboratory evaluation, and storage of biological specimens for genetic or molecular analysis in this trial/future use

Fecal swabs and stool samples will be collected by the participants themselves before the study intervention begins. Instructions and necessary materials for the collections will be sent to the participant’s address. After collecting the samples, the participant should coordinate their delivery with the investigators. The biological samples processing will be conducted at BiomeHub. Fecal swab samples will be used to assess the volunteer’s gut microbiome by metagenomics shotgun technique, and stool samples will be used for microorganism isolation. As for the biological samples collected, they will be stored until they are processed, and after that, they will be discarded. The raw data from bioinformatics analyses will be stored for up to five years after the completion of the study, by current laws and regulations.

## Statistical method

### Statistical methods for primary and secondary outcomes

Following a pre-specified statistical analysis plan (Supplementary data), the primary objectives will be analyzed using random effects linear models with Gaussian likelihood. Random effects will vary by participant and include an intercept and a slope coefficient for the diet effect. A fixed interaction between the diet effect and a pre-specified gut microbiome score will also be computed. Likelihood ratio tests will be employed at a significance level of 5%. We will correct p-values for multiple comparisons using the Homl’s method due to the two co-primary objectives. Each participant in the study will have an estimate of their intrinsic responsiveness to the tested diets (i.e., the individual treatment effect of the comparator diet). Sensitivity analyses will be implemented to check the robustness of the inferences made during the primary analysis, including period-by-treatment interactions, smooth time trends using restricted cubic splines, and adjustment by randomized sequence.

All analyses will be conducted using the R software package (version 4.3.0 or higher) ^25^. A fixed Docker image will be used for all analyses to ensure full reproducibility ^26^. The simulation-based power calculation performed for this document is available at https://github.com/biomehub/bhub-n-of-1-sap and is equally fully reproducible.

### Interim analyses

No interim analyses are planned for this trial. The final analysis will be conducted once the number of recruited patients reaches the specified sample size required for the study.

### Methods for additional analyses (e.g. subgroup analyses)

Secondary analyses will further explore the influence of the gut microbiome profile on the individual treatment effects of Diet B on the iAUC. In particular, the relationship between the microbiome-by-treatment interaction will allow for non-linearities using restricted cubic splines. The conditional effect of diet B given the microbiome score will then be assessed visually. Exploratory microbiome data analyses will also be implemented. Associations between new biomarkers and the individual treatment effects of diet B will be analyzed similarly to the primary analysis, replacing the patients’ gut microbiome score values with the corresponding biomarker values. Given its exploratory nature, this analysis will control the false discovery rate (FDR) at 10% using the Benjamini-Hochberg procedure ^27^.

### Methods in analysis to handle protocol non-adherence and any statistical methods to handle missing data

If observed, missing covariate data will be handled using Multivariate Imputation by Chained Equations. Sensitivity analyses will include a complete case analysis and a corresponding Bayesian analysis treating missing data as parameters. The random effects model for the primary analysis will directly handle missing outcome data, under the assumption that missingness happens at random.

### Plans to give access to the full protocol, participant-level data, and statistical code

After the end of the study and analyses, participants will have access to their results. The project’s results, anonymised participant dataset, will become publicly available through article publications. If any researcher has any questions about the protocol or the publications, they can contact the corresponding researcher.

## Oversight and monitoring

### Composition of the coordinating center and trial steering committee

Organizational structure and responsibilities: BiomeHub will act as the study sponsor taking all responsibility of study conduct. The principal investigator is responsible for study design and conduct, preparation of protocol and revisions, organizing steering committee meetings, publication of study reports and members of the Trial *Management Committee*.

Trial Management Committee (TMC) will be formed by principal investigator and coinvestigators that will be responsible for study planning, organization of steering committee meetings, responsible for the trial master file, budget administration, data verification, randomization, organization of sample collection.

Data Manager will be represented by principal investigator and coinvestigators and will be responsible for the maintenance of the trial IT system and data entry, and data verification.

### Composition of the data monitoring committee, its role, and reporting structure

No data monitoring committee was assigned for this single-center trial without major safety concerns. Any patient-reported discomfort will be monitored by study investigators, who will have access to all data throughout.

### Adverse event reporting and harms

The risks associated with participation in this research are low. The collection of fecal samples does not cause any pain or discomfort to the participant; it may only be an unpleasant procedure. The insertion of the continuous glucose monitoring sensor, performed using a specific applicator that makes a superficial skin puncture (5mm deep), can cause some discomfort. Additionally, the presence of the device may generate discomfort during use if there are accidental situations, such as hitting the area where the device is inserted or applying pressure to the device on the skin. Participants may feel uncomfortable when responding to the study-related questionnaire, which includes questions about their personal and clinical data. However, the project’s researchers will always be available to clarify any doubts and ensure that participants feel comfortable providing this information. If participants feel that they have been harmed or experience any issues related to the study, they can immediately report the problem via email to the responsible researchers.

### Frequency and plans for auditing trial conduct

There will not have formal auditing of the trial by the investigating team.

### Plans for communicating necessary protocol amendments to relevant parties (e.g. trial participants, ethical committees)

There is no plan to modify the study protocol or interventions. However, if there are any changes to the study design, eligibility criteria, or sample sizes that can impact study conduct will initially require sponsor and investigators agreement and then, they will be reported to the Ethics Committee for Research. If necessary, the participants will be notified of the changes and will sign an updated informed consent form with the modifications.

### Dissemination plans

The study is reported in Clinical Trials Registration by NCT06051318 number. Study protocol abstracts and preliminary results may be submitted for a presentation to relevant scientific symposia and conferences. The trial results will be published in a peer-reviewed journal. Trial results will be reported to study collaborators and participants following study completion.

### Discussion

The present study represents a step forward in understanding the relationship between the intestinal microbiome and glycemia. Several studies have addressed the potential of the microbiome to predict glycemic response, including observational cohort studies, non-randomized or single-arm experimental trials, and randomized parallel or crossover trials ^28–31^. The main objective shared by these studies is to use the microbiome to guide personalized nutritional interventions ^32^. Their shared limitation, however, is the inability to separate variation of individual glycemic responses from causal treatment effects at the individual level ^33^.

For any given patient, being able to estimate individual treatment effects (ITE) relies on her/his repeated exposure to different treatments, in a design known as N-of-1 trial ^34^. In the present series of N-of-1 trials, our objective is twofold: (A) assess heterogeneity in the ITE of diet on glycemic response and (B) investigate the relevance of the intestinal microbiome profile as a predictor of this heterogeneity. The first objective addresses the often overlooked need to ensure variation across individuals pertains not only observed responses and their associations but also truly causal effects suitable to inform decision making. The second objective is essential to identify predictive biomarkers, which modify the expected treatment effect of the studied diets, going beyond pure associations with glycemic response. Therefore, while objective (A) addresses the underlying assumption behind the hope for personalized dietary interventions, objective (B) investigates the necessary tools to identify those who intrinsically benefit the most from which diet, outside the trial population. As our study is particularly well-powered, the eventual failure to observe significant variations of ITE casts major doubts onto the underlying principle of precision nutrition ^35^.

In the future, we hope to continue refining a programmatic template for using personalized N-of-1 designs for health conditions, such as metabolic diseases, and other interventions, such as another type of diet. Furthermore, the results of this study are expected to assist healthcare professionals in managing personalized diets for glycemic control. Understanding the relationships between the intestinal microbiome, individual characteristics, and glycemic response can enable healthcare professionals to develop more precise and effective nutritional strategies for patients with diabetes, pre-diabetes, and other glycemic-related conditions.

### Trial status

Protocol version: 1

Start date of the recruitment: January 2024

Recruitment final date: June 2024

## Data Availability

All data produced in the present study are available upon reasonable request to the authors

## Abbreviations

CGM: Continuous Glucose Monitor
FDR: False Discovery Rate
ITE: Individual treatment effects
TMC: Trial Management Committee
iAUC: incremental Area-Under-the-Curve

## Acknowledgments

We would like to acknowledge other participating researchers involved in the project: Aline F. R. Sereia (BiomeHub), Ana Paula Christoff (BiomeHub), Daniela C. De Bastiani (BiomeHub), Michele P. Rode (BiomeHub), Milene H. de Moraes (BiomeHub), Natália M. Gutierrez (BiomeHub). We would like to thank the nutritionists Fernanda Piazza (BiomeHub), Louise Crovesy de Oliveira (BiomeHub) and Sabrina Reis (BiomeHub), who designed the two standardized meals that will be distributed in this study.

## Authors’ contributions {31b}

LFO is the sponsor of this study. The study idea and concept were developed by LFO, CPZ, BLT and GNFC. The draft of this study was prepared by investigators CPZ, BLT and GNFC who also revised and approved the final version in collaboration with the principal investigator. CPZ, BLT, VBF and GNFC will be responsible for the data analysis and for writing the manuscript with final results.

## Funding

This study will be funded by the biotechnology company BiomeHub and partially supported by the National Committee for Scientific and Technological Development (CNPq) with two scholarships from the Human Resources Training Program in Strategic Areas (RHAE), process number 424464/2021-7.

## Availability of data and materials

All investigators will be given access to the cleaned data sets. The project data will be stored in a folder created for the study, and access will be limited to only the investigators. Investigators will have direct access to their own site’s data sets, and will have access to other sites’ data by request. To ensure confidentiality, data dispersed to project team members will be blinded to any identifying participant information.

## Ethics approval and consent to participate

This study has been approved by the National Committee for Research Ethics (CONEP) under the number 6.225.662. Informed consent forms will be collected from all participants before they begin the study.

## Competing interests

The authors declared the following potential conflicts of interest concerning the research, authorship and publication of this protocol: Luiz Felipe Valter de Oliveira is the BiomeHub Technologies startup CEO, in which the project was designed and will be conducted. Bianca Luise Teixeira and Giuliano Netto Flores Cruz are employees at BiomeHub and will actively work on the project. Caetana Paes Zamparette and Vilmar Benetti Filho receive CNPq scholarships for the development of this project at BiomeHub and will actively contribute to the project.

## Notes

### Clinical Trial

NCT06051318

### Author Declarations

Committee on Ethics in Research - Center for Oncological Research -CEPON Conclusions or Pending and List of Inadequacies: All the pending issues presented in the Substantiated Opinion CEP No. 6,112,074, dated June 12, 2023, have been duly addressed and approved.

