## Supplemental data for "Individual effect of diet on postprandial glycemic response and its relationship with gut microbiome profile in healthy subjects: protocol for a series of randomized N-of-1 trials"

### Statistical Analysis Plan (SAP)

#### Gut microbiome and individual treatment effects of diet on postprandial glucose: a series of N-of-1 trials

Version 1.0.0

BiomeHub
Department of Bioinformatics

21st December, 2023

**5 Software and reproducibility** **7**

### 1 Administrative information

The history for the current Statistical Analysis Plan (SAP) is shown in Table 1. The current SAP version (1.0.0) is aligned with protocol version 1.0.0. Trial registration: NCT06051318.

Table 1: SAP history.

| Protocol version | Updated SAP version | Section | Description | Date |
|---|---|---|---|---|
| 1.0.0 | 1.0.0 | — | Initial version | 21/Dec/2023 |

#### 1.1 Roles and responsibility

This SAP was written by Giuliano Netto Flores Cruz, who is responsible for the statistical design and analysis of the trial. The SAP was reviewed and approved by all investigators.

### 2 Study design

#### 2.1 Design overview

This is a series of N-of-1 trials comparing two diets (breakfast meals). The reference treatment is denoted Diet A, while the comparator is Diet B. The primary outcome is the positive incremental area-under-the-curve (iAUC) of the postprandial blood glucose, measured in $\text{mmol} \cdot \text{min} \cdot \text{L}^{-1}$ and calculated according to Brouns et al. (2005) [1]. The iAUC is based on continuous glucose monitoring with a blood glucose measurement right before and every 30 minutes for 2 hours after breakfast. Within each of five treatment cycles, each participant undergoes two periods of treatment. Within each of the ten total treatment periods, one diet is consumed and the outcome is measured. Participants will be randomized into one of two doubly counterbalanced sequences: ABBABAABBA or BAABABBAAB [2, 3]. Analysis of all outcomes will be carried out at the end of the study when all relevant data is available.

#### 2.2 Data generating process

Here we state the full specification of the data generating process used for simulation-based power calculations. It also lays the basis for the primary analysis plan, described in upcoming sections.

Let $Y_{ij}$ be the iAUC for participant $i = 1, 2, \ldots, n$ measured at the period $j = 1, 2, \ldots, 10$. Let $X_{ij}$ be the corresponding treatment indicator variable – i.e., it takes the value of 1 if the $i^{\text{th}}$ participant ate diet B in the $j^{\text{th}}$ period, and 0 otherwise. Denote $M_i$ as a continuous score representing the gut microbiome profile for the $i^{\text{th}}$ participant at the start of the trial (centred and scaled to variance 1). Also, define $S_i$ as the indicator variable for the treatment sequence randomized to each participant (equals 1 if the sequence is BAABABBAAB and 0 otherwise). We then assume

$$Y_{ij} = \alpha_i + \beta_i X_{ij} + g(j) + \kappa S_i + \epsilon_{ij} \tag{1}$$

where $g(j) = \sum_{t=2}^{5} \mu_t \cdot \mathbb{1}\{t = j\}$ is the effect of the $j^{\text{th}}$ period, for $j = 2, \ldots, 10$. Due to randomization, we know that the average sequence effect $\kappa$ is exactly zero, but it is included to increase precision.

For the $i^{\text{th}}$ participant, $\alpha_i$ is their average iAUC under Diet A and $\beta_i$ is the average individual effect of Diet B. Note that $\beta_i$ is an Individual Treatment Effect (ITE), identifiable thanks to the N-of-1 design where each individual participant, considered as the population of interest, is treated multiple times. The independent error term is $\epsilon_{ij} \sim \mathcal{N}(0, \sigma_\epsilon^2)$ and represents the residual variability within participant and period. The participant-specific terms are modeled as

$$\alpha_i = \alpha_0 + \tau_1 M_i + u_{1i} \tag{2}$$

$$\beta_i = \beta_0 + \tau_2 M_i + u_{2i} \tag{3}$$

where $u_{1i}$ and $u_{2i}$ are random intercept and random slope terms, respectively, given by

$$\begin{pmatrix} u_{1i} \\ u_{2i} \end{pmatrix} \sim \mathcal{N}\Big(\mathbf{0}, \mathbf{\Sigma}\Big) \tag{4}$$

$$\mathbf{\Sigma} = \begin{pmatrix} \sigma_1^2 & \sigma_{12} \\ \sigma_{12} & \sigma_2^2 \end{pmatrix} \tag{5}$$

Table 2 shows the interpretation for each parameter in the data-generating process specified by (1) — (5) along with values used in the simulation-based power calculations (see below). In addition to the ITE, $\beta_i$, we can also estimate the overall average treatment effect (ATE) through $\beta_0$. The interaction term $\tau_2$ allows estimating the conditional average treatment effect (CATE) of diet B, given the microbiome profile $M_i$.

Table 2: Parameters of the assumed data-generating process specified by (1)—(5).

| Parameter | Interpretation | Simulated value |
|---|---|---|
| $\alpha_i$ | Average iAUC for the $i^{\text{th}}$ individual eating diet A with average microbiome profile (i.e., $M_i = 0$) | † |
| $\beta_i$ | Average individual treatment effect (ITE) of diet B on iAUC for the $i^{\text{th}}$ individual | † |
| $g(j)$ | Average effect of $j^{\text{th}}$ period (period $j = 1$ is the reference) | 0 |
| $\kappa$ | Average effect of treatment sequence (sequence ABBABAABBA is the reference), known to be zero due to randomization | 0 |
| $\alpha_0$ | Overall average iAUC under Diet A | 200 |
| $\tau_1$ | Average effect on iAUC of increasing $M_i$ by 1 standard deviation | 5 |
| $\beta_0$ | Overall average treatment effect (ATE) of diet B on iAUC | 0 |
| $\tau_2$ | Conditional average treatment effect (CATE) of diet B on iAUC representing microbiome-by-treatment interaction | 5 |
| $\sigma_1^2$ | Between-patient variance of $u_{1i}$ (larger values represent greater variability of average iAUCs across individuals) | 1575 |
| $\sigma_2^2$ | Between-patient variance of $u_{2i}$ (larger values represent stronger patient-by-treatment interaction, i.e., greater variability of ITEs) | 75 |
| $\sigma_{12}$ | Covariance between $u_{1i}$ and $u_{2i}$ | 0 |
| $\sigma_\epsilon^2$ | Residual within-patient variance (how much iAUC varies within a given patient, from period to period, under the same treatment) | 225 |

† These values are computed from other parameters.

The rationale for the parameter values used in Table 2 was as follows. From past literature, it was assumed that 99% of potential iAUC values range approximately between 100 and 300 $\text{mmol} \cdot \text{min} \cdot \text{L}^{-1}$, with an average of 200 $\text{mmol} \cdot \text{min} \cdot \text{L}^{-1}$ [4, 5, 6]. A relative difference in iAUC of 20% was used to compute the minimal clinically important difference (MCID) [7, 8, 9]. With a baseline average iAUC of 200 $\text{mmol} \cdot \text{min} \cdot \text{L}^{-1}$, this corresponds to an absolute MCID of 40 $\text{mmol} \cdot \text{min} \cdot \text{L}^{-1}$. In light of this value, the parameters $\sigma_2^2$ and $\tau_2$ were set such that $\beta_i$ would be within $\beta_0 \pm 20$ with approximately 95% probability (i.e., the standard deviation of $\beta_i$ was set to 10). This means that the expected difference between the highest individual effect and the lowest individual effect should be close to 40 $\text{mmol} \cdot \text{min} \cdot \text{L}^{-1}$ around 95% of the time. The value of $\tau_2$ was chosen so that around 25% of the total variance of the ITEs ($\beta_i$) was attributable to the microbiome score $M_i$. Similarly, the individual average iAUCs ($\alpha_i$) were assumed to lie in the range 120—280 $\text{mmol} \cdot \text{min} \cdot \text{L}^{-1}$ about 95% of the time, with 25% of this variation attributable to variation in the microbiome, which defined the values of $\sigma_1^2$ and $\tau_1$ shown in Table 2. Finally, we assumed that the *observed* iAUC values for each patient, under a given treatment, vary from period to period within $\pm 30$ $\text{mmol} \cdot \text{min} \cdot \text{L}^{-1}$ of their individual averages.

### 3 Primary analysis plan

The primary analysis plan will be based on random effects models and likelihood-ratio tests at a 5% significance level, described below for the two co-primary objectives of the study. We will employ Holm's method to correct p-values due to the multiple comparisons. Missing covariate data will be handled using Multiple Imputation. The random effects models naturally deal with missing outcome values (e.g., if there is no iAUC available in some treatment period), assuming these are missing at random. Any estimates will be reported alongside 95% confidence intervals (two-sided) and will be considered in light of the MCID of 40 $\text{mmol} \cdot \text{min} \cdot \text{L}^{-1}$.

#### 3.1 Determining the presence of heterogeneity of the ITEs of Diet B on iAUC — patient-by-treatment interaction

In light of the data generating process defined by (1)—(5), here the specific question is: does $\beta_i$ vary significantly across participants? This is equivalent to testing the (random) patient-by-treatment interaction. Notice that the influence of the microbiome profile $M_i$ does not matter for this question.

The full model is:

$$Y_{ij} = \alpha_0 + u_{1i} + (\beta_0 + u_{2i})X_{ij} + g(j) + \kappa S_i + \epsilon_{ij} \tag{6}$$

The corresponding reduced model is:

$$Y_{ij} = \alpha_0 + u_{1i} + \beta_0 X_{ij} + g(j) + \kappa S_i + \epsilon_{ij} \tag{7}$$

In other words, we will test whether the use of a random slope term $u_{2i}$ improves the model fit due to significant variation in the effect of Diet B from participant to participant – i.e., the patient-by-treatment interaction.

#### 3.2 Determining the influence of the gut microbiome on the heterogeneity of the ITEs — microbiome-by-treatment interaction

Here the specific question is: does $\beta_i$ vary significantly with $M_i$? This is equivalent to testing the (fixed) microbiome-by-treatment interaction. If indeed $\beta_i$ varies across participants, then we may be able to explain some of this variation by conditioning on the microbiome profile score $M_i$. This conditional average treatment effect (CATE) of Diet B given microbiome profile $M_i$ can then be used to inform decision-making for future patients, outside the trial.

The full model is:

$$Y_{ij} = \alpha_0 + \tau_1 M_i + u_{1i} + (\beta_0 + \tau_2 M_i + u_{2i})X_{ij} + g(j) + \kappa S_i + \epsilon_{ij} \tag{8}$$

The corresponding reduced model is:

$$Y_{ij} = \alpha_0 + \tau_1 M_i + u_{1i} + (\beta_0 + u_{2i})X_{ij} + g(j) + \kappa S_i + \epsilon_{ij} \tag{9}$$

So here we are testing whether $\tau_2 = 0$ — i.e, the microbiome-by-treatment interaction. Notice that Eq. (8) is exactly Eq. (1). The underlying linearity assumption between $M_i$ and $\beta_i$ will be relaxed in secondary analyses when estimating the effect of Diet B given an observed microbiome profile score $M_i$ (e.g., using restricted cubic splines). The full model given by Eq. (8) will be used to estimate ITEs, regardless of statistical significance. ITEs will be reported as point estimates and bootstrap-based 95% confidence intervals.

#### 3.3 Sample size and statistical power

The sample size was determined by simulation-based power calculation, under the assumptions described in section 2.2. The simulation setting followed the full data generating process described by Eq. (1)—(5) with parameters defined in Table 2. Using likelihood ratio tests at a 5% significance level and assuming a dropout rate of 20%, the present study required the recruitment of 80 participants followed through five treatment cycles. The numbers of participants and of treatment cycles were selected to provide a statistical power of at least 90% and 85% for the detection of patient-by-treatment interaction and microbiome-by-treatment interaction, respectively. The power calculation considered p-values adjusted for multiple comparisons using Holm's method. Given these assumptions and the primary analysis plan described above, the present study required 80 participants with 5 treatment cycles per participant.

#### 3.4 Sensitivity analyses

Sensitivity analyses will be implemented to check the robustness of the inferences made during the primary analysis, including period-by-treatment interactions and smooth time trends using restricted cubic splines. If some covariate data is missing (e.g., missing $M_i$), we will also perform a complete case analysis as well as a Bayesian version of the primary analysis treating each missing data point as a parameter. A Bayesian version of the primary analysis will also be performed, using weakly- or non-informative priors. The Bayesian interpretation of the results will be reported as a supplement.

### 4 Secondary analyses plan

#### 4.1 Non-linear influence of the gut microbiome score on the heterogeneity of the ITEs

Secondary analyses will further explore the influence of the gut microbiome profile on the individual treatment effects of Diet B on the iAUC. In particular, the relationship between the treatment-by-microbiome interaction will allow for non-linearities using restricted cubic splines. The conditional effect of diet B given the microbiome score will be assessed visually.

#### 4.2 Exploratory microbiome data analysis

Exploratory microbiome data analyses will be implemented [10]. Associations between new biomarkers and the individual treatment effects of diet B will be analyzed similarly to the primary analysis described in section 3.2, replacing the gut microbiome score $M_i$ with the $i^{\text{th}}$ patient's corresponding biomarker value. These biomarkers will include the relative abundance of the observed taxa (at the oligotype, species, genus, family, and phylum levels), alpha-diversity metrics (Shannon and Inverse Simpson indexes), functional microbiome profiles, and biochemical and inflammatory markers. Using proportion-normalized oligotype abundances, we will also perform beta-diversity analysis (e.g., based on Bray-Curtis dissimilarity). Differential abundance (DA) analyses will be performed as appropriate, considering the consensus of at least two DA tools available [11].

Given its exploratory nature, this analysis will control the false discovery rate (FDR) at 10% using the Benjamini-Hochberg procedure [12]. Potential predictive models for both individual treatment effects and individual response will be explored using machine learning methods and internally validated through cross-validation. The predictive models will be $\ell_2$- and/or $\ell_1$-penalized for complexity to mitigate overfitting.

### 5 Software and reproducibility

All analyses will be conducted using the R software package (version 4.3.0 or higher) with code versioning and software dependency tracking [13]. A fixed Docker image will be used for all analyses to ensure full reproducibility [14]. Random effects models will be estimated using the lme4 R package (v. 1.1.33 or higher) and general data analysis will employ the tidyverse R meta-package [15, 16]. The simulation-based power calculation performed for this document is available at https://github.com/biomehub/bhub-n-of-1-sap and is equally fully-reproducible.

[10] Rob Knight, Alison Vrbanac, Bryn C. Taylor, Alexander Aksenov, Chris Callewaert, Justine Debelius, Antonio Gonzalez, Tomasz Kosciolek, Laura-Isobel McCall, Daniel McDonald, Alexey V. Melnik, James T. Morton, Jose Navas, Robert A. Quinn, Jon G. Sanders, Austin D. Swafford, Luke R. Thompson, Anupriya Tripathi, Zhenjiang Z. Xu, Jesse R. Zaneveld, Qiyun Zhu, J. Gregory Caporaso, and Pieter C. Dorrestein. Best practices for analysing microbiomes. *Nature Reviews Microbiology*, 16(7):410–422, May 2018. ISSN 1740-1534. doi: 10.1038/s41579-018-0029-9. URL http://dx.doi.org/10.1038/s41579-018-0029-9.

[16] Hadley Wickham, Mara Averick, Jennifer Bryan, Winston Chang, Lucy McGowan, Romain François, Garrett Grolemund, Alex Hayes, Lionel Henry, Jim Hester, Max Kuhn, Thomas Pedersen, Evan Miller, Stephan Bache, Kirill Müller, Jeroen Ooms, David Robinson, Dana Seidel, Vitalie Spinu, Kohske Takahashi, Davis Vaughan, Claus Wilke, Kara Woo, and Hiroaki

Yutani. Welcome to the tidyverse. *Journal of Open Source Software*, 4(43):1686, November 2019. ISSN 2475-9066. doi: 10.21105/joss.01686. URL http://dx.doi.org/10.21105/joss.01686.
